# Serum RNA profiling in the 10-year period prior to diagnosis of testicular germ cell tumour

**DOI:** 10.1101/2020.06.03.20121087

**Authors:** Joshua Burton, Sinan U. Umu, Hilde Langseth, Tom Grotmol, Tom K. Grimsrud, Trine B. Haugen, Trine B. Rounge

## Abstract

Although testicular germ cell tumour (TGCT) overall is highly curable, patients may experience late effects after treatment. An increased understanding of the mechanisms behind the development of TGCT may pave the way for better outcome for patients. To elucidate molecular changes prior to TGCT diagnosis we sequenced small RNAs in serum from 69 patients who were later diagnosed with TGCT and 111 matched controls. The deep RNA profiles, with on average 18 million sequences per sample, comprised of nine classes of RNA, including microRNA. We found that circulating RNA signals differed significantly between cases and controls regardless of time to diagnosis. Different levels of TSIX related to X-chromosome inactivation and TEX101 involved in spermatozoa production are among the interesting findings. The RNA signals differed between seminoma and nonseminoma TGCT subtypes, with seminoma cases showing lower levels of RNAs and nonseminoma cases showing higher levels of RNAs, compared with controls. The differentially expressed RNAs were typically associated with cancer related pathways. Our results indicate that circulating RNA profiles change during TGCT development according to histology and may be useful for early detection of this tumour type.

## Introduction

Testicular germ cell tumour (TGCT) is diagnosed in 1% of men worldwide and is the most common malignancy in males between 20 and 39 years of age. The incidence rates are still rising [Trabert, 2015], and some of the highest incidences are found in Northern Europe [Znaor et al., 2014]. The etiology is largely unknown, although genetic components and conditions during pregnancy seem to play a role. [Elzinga-Tinke et al., 2015]. The susceptibility to TGCT is shown to have a strong familial link, with a fourfold increased risk for fathers and eightfold for brothers [Hemminki and Li, 2004; Dong et al., 2001]. The polygenic nature of TGCT has been recognised and more than 50 susceptibility genes have been identified [Kristiansen et al., 2015; Wang et al., 2017; Litchfield et al., 2018; Das et al., 2019]. The susceptibility loci contain genes linked to germ cell development and sex determination, as well as genes related to tumour growth/suppression. The main histologic subtypes of TGCT are seminoma, nonseminoma and mixed, making up around 50%, 30% and 20%, respectively [Oosterhuis and Looijenga, 2005].

TGCT is a highly curable disease since the introduction of cisplatin-based chemotherapy with a 5-year survival rate of above 95% [Trama et al., 2015]. However, there is an increased risk of long-term side effects including secondary non-germ cell (GC) cancers, cardiovascular disease, and hypogonadism. [Haugnes et al., 2012; Raphael et al., 2019; Bucher-Johannesen et al., 2019].

The current guidelines presented by the European Association of Urology [Laguna, 2020] for the diagnosis of TGCT include several techniques, including ultrasound imaging for initial diagnosis and serum markers for specific subtype diagnosis and prognosis [Richie et al., 1982; Germà-Lluch et al., 2002: Murray et al., 2016]. The serum markers used are alpha-fetoprotein, hCG and LDH, however alpha-fetoprotein is only seen in yolk-sac tumour and hCG is expressed by trophoblasts only. The presence of histological markers is also used for diagnosis. For germ cell neoplasia *in situ* (GCNIS), the markers include OCT3/4, PLAP, CD117, and SALL 4 [Laguna, 2020]. Furthermore, it has been shown that the metabolic biomarkers leptin and resistin may predict cancer mortality due to their function of inducing pro-tumorigenic environment that further promotes tumour initiation, angiogenesis, and metastasis [Akinyemiju, 2018].

RNAs, such as microRNA (miRNA), piwi-interacting RNA (piRNA), and long non-coding RNA (lncRNA), regulate gene expression on transcriptional and post-transcriptional levels and have been shown to be present in serum [Umu et al., 2018]. The differential composition of RNAs in circulation can help determine diagnosis as well as the developmental stage of the tumour [Fernandez-Mercado et al., 2015]. Circulating miRNAs have been identified as both prognostic and diagnostic markers in biliary tract cancer (BTC), which has led to earlier diagnosis and less invasive procedure compared with previously used techniques [Letelier, 2016]. Exosomal miRNAs have also been observed in colorectal cancer (CRC). Specifically, miR-150-5p and miR-99b-5p were found to be downregulated in CRC patients compared to healthy patients [Zhao et al., 2019]. Differential levels of circulating RNA were also observed in pre-diagnostic serum samples in lung cancer patients, with an overall dynamic trend when advancing clinically [Umu et al., 2020].

In TGCT patients, high expression of miRNAs belonging to the clusters *miR-302/367* and *miR-371-373* has been found in serum [Palmer et al., 2010; Murray et al., 2011; Gillis et al., 2007]. Small RNA sequencing performed on TGCT tissue samples revealed miRNAs profiles to differ between normal and TGCT tissues, as well as between histological subtypes. A genome wide downregulation or loss of piRNAs was observed in TGCT, through mechanisms such as hypermethylation in CpG islands on genes associated with piRNAs [Rounge et al., 2015; Ferreira et al., 2014; Das et al., 2019].

Circulating RNAs in TGCT patients have allowed for a less invasive and a more specific histology at diagnosis [Laguna, 2020]. However, pre-diagnostic circulating RNA changes in TGCT have yet to be studied.

The aim of this study was to investigate the role of RNA in the development of TGCT in serum samples collected in a 10-year period prior to diagnosis. Furthermore, we investigated how differences in circulating RNAs were related to histologies and time periods before diagnosis. Differences in RNAs profiles were then used in functional enrichment analysis, and RNAs with stable patterns were investigated further.

## Materials and Methods

### Study design and participants

All samples included were retrieved from the Janus Serum Bank (JSB). JSB is a population-based cancer research biobank containing serum samples from 318 628 individuals collected from 1972 to 2004 [Langseth et al., 2017; Hjerkind et al., 2017]. The TGCT cases were identified by linking the JSB to the Cancer Registry of Norway using the Norwegian individually unique national identity numbers. The prediagnostic serum samples were donated up to 10 years before the diagnosis. We drew 111 cancer-free Janus participants for comparison of RNA levels with the cancer cases. The control subjects had to be alive and free from cancer, except for non-melanoma skin cancer, at the time of their matched case’s diagnosis, and up to 10 years after blood collection. Controls were matched on age, time of blood collection and blood donor group (dependent on their county of residence, year of collection and method of storage), and as a result of these matching criteria, the average age at sample donation was 35 years (standard deviation of 6.5/6.7 respectively) for both cases and controls.

Previous studies have shown the feasibility of using long-term archived serum samples for RNA analyses and the variability of the RNA levels [Umu, et al., 2018; Rounge et al., 2015; Rounge et al., 2018]. The methods and analyses of the TGCT serum samples were similar to studies by Umu et al [Umu et al., 2019]. The donors have given broad consent for the use of the samples in cancer research. The study was approved by the Norwegian regional committee for medical and health research ethics (REC no: 24 846, 2012/1590).

### Laboratory processing of serum to RNA profiles

We extracted RNAs from 400 μl serum using phenol-chloroform phase separation and the miRNeasy Serum/Plasma kit (Cat. no 1071073, Qiagen) using a QIAcube (Qiagen). NEBNext® Small RNA Library Prep Set for Illumina (Cat. No E7300, New England Biolabs Inc.) was used for small RNA-seq library preparation. RNA molecules from 17 to 47 nt in length were selected. We sequenced 12 samples per lane of a HiSeq 2500 (Illumina). Additional information is available in our previous study [Umu et al., 2018].

### Bioinformatics analyses

Total number of reads generated was 3.5 billion with an average sampling depth of 18.4 million raw reads. AdapterRemoval v2.1.7 [Schubert et al., 2016] was used to trim for adapters. We then mapped the collapsed reads to human genome version hg38 with Bowtie2 v2.2.9, with 10 alignments per read being allowed. Our annotation set consist of miRBase(v.22) [Kozomara and Griffiths-Jones, 2014] for miRNAs, pirBAse for piRNAs [Zhang et al., 2014] and GENCODE [Harrow et al., 2012] for other RNAs. IsomiR and tRF profiles were obtained through SeqBuster [Pantano et al., 2010] and MINTmap, respectively [Loher et al., 2017]. In the analyses, we included RNAs with at least 5 reads in more than 20% of the samples. Details are available in our previous study [Umu et al., 2018].

The *optmatch* R package [github.com/markmfredrickson/optmatch] allowed us to find optimally matched sets of controls (Table S1.) for each analysis. The analyses were matched on age, histology when appropriate and technical artefacts. The technical artefacts accounted for differences in pre-analytical treatment and storage time [Rounge et al., 2015; Umu et al., 2018], termed blood donor groups. The DESeq2 R package (v1.18.1) [Love et al., 2014] was used for the differential expression analyses using the default generalized linear model with a negative binomial distribution.

Kyoto Encyclopedia of Genes and Genomes (KEGG) pathway analysis was performed using kegga function from the limma R package. The inputs included mRNA, miRNA and isomiR targets extracted from miRDB (v5.0) predictions [Wong and Wang, 2015] with a cut off score of >60.

To exploit the full statistical power of the dataset when identifying RNAs that differ in cases and controls regardless of other factors, we first compared all cases and controls. To identify histology (determined by ICD-O (3rd revision) codes) specific RNA signals, we compared the RNA levels of seminoma with those of the matched controls, and similarly for nonseminomas and matched controls. To identify RNA signals according to the time between blood draw and diagnosis we divided prediagnostic time into four discrete time intervals, 0 to 2, 2 to 5, 5 to 8 and 8 to 10 years. We selected these intervals to optimize resolution on time to diagnosis while still having sufficient statistical power. To make the time windows comparable with respect to statistical power, the cut points were chosen to secure the same number of cases and controls, and similar proportions of histologies.

## Results

### Reduced levels of RNAs in TGCT cases

To identify differentially expressed RNAs in all TGCT cases (n=79) against matched controls (n=111), we analysed 4231 RNAs that passed our inclusion criteria. Of these, we identified 818 RNAs that were differentially expressed with a p-adjusted value ≤ 0.05, and 88 of these had a log2 fold change (log2fc) outside the range {-1,1} (figure 1a.). The majority of these RNAs (82) had reduced levels in cases compared to controls, with an average log2fc = - 1.37 and p-adjusted = 0.00032. The RNAs showing reduced levels were primarily isomiRs and mRNA fragments, whereas the few elevated RNAs consisted of lncRNAs, mRNA fragments and a tRF (figure 1b.).

**Figure 1.**
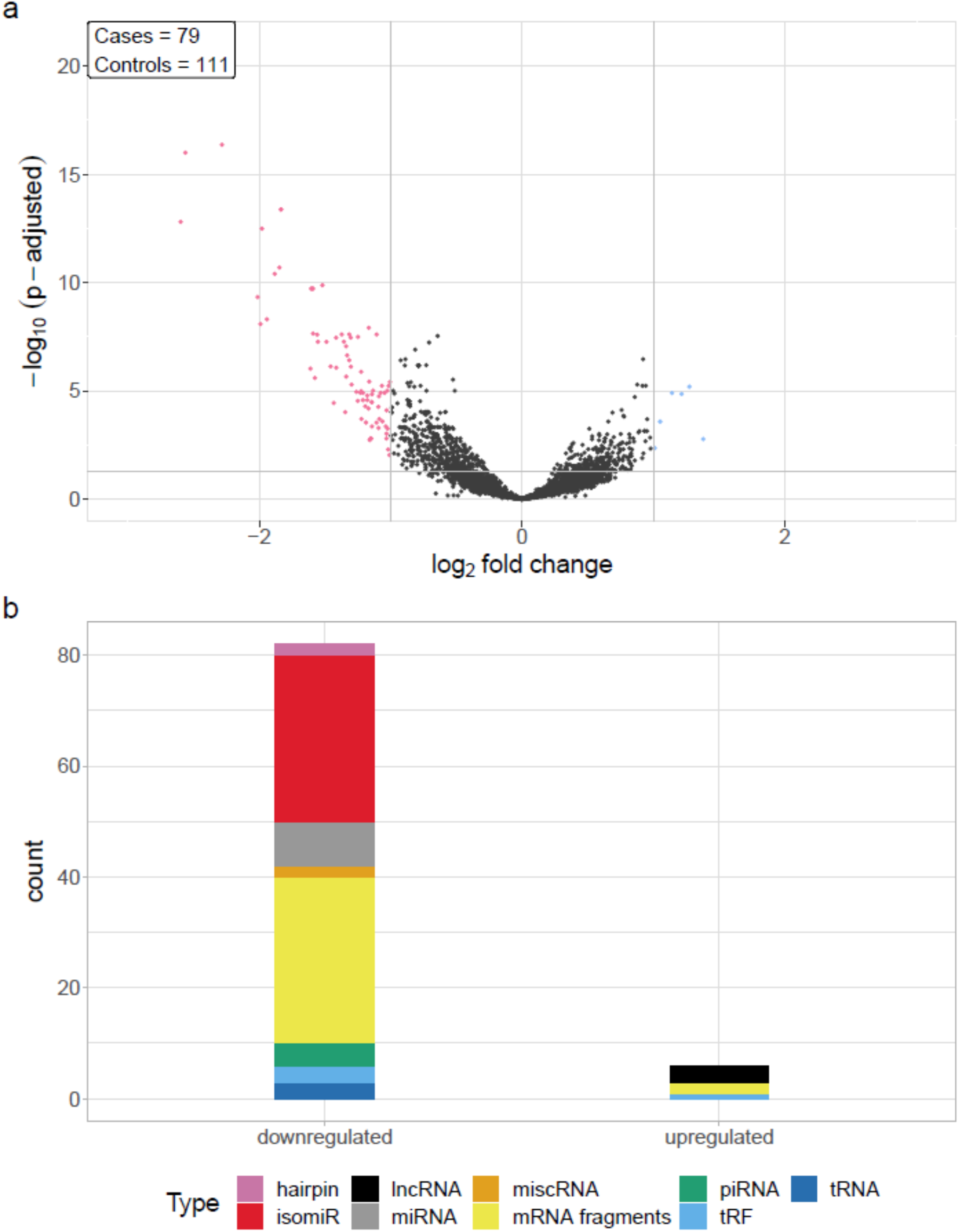
a) Differential RNA levels in pre-diagnostic serum samples from 79 TGCT cases and 111 matched controls. Pink represent RNAs with >1 log2 fold change reduced levels in cases and blue represent RNAs with >1 log2 fold change increased levels in cases, compared to controls. b) RNA class composition of the significantly elevated and reduced RNAs.

### RNA signals differ with histological subtype

We compared RNA levels in seminoma and nonseminoma cases with control samples. As seen for all cases combined, seminomas exhibited a similar pattern with reduced level for the majority of the differentially expressed RNAs.

In total, 112 RNA signals were different in seminomas compared to controls, 102 with lower RNA levels and 10 with elevated levels (figure 2a). Similar to all TGCT cases, the majority of the downregulated RNAs, with a mean log2fc values of -1.35, consisted of mRNA fragments and isomiRs, whereas the upregulated RNAs consisted of lncRNAs, mRNA fragments, piRNAs, and tRFs (figure 2c).

**Figure 2.**
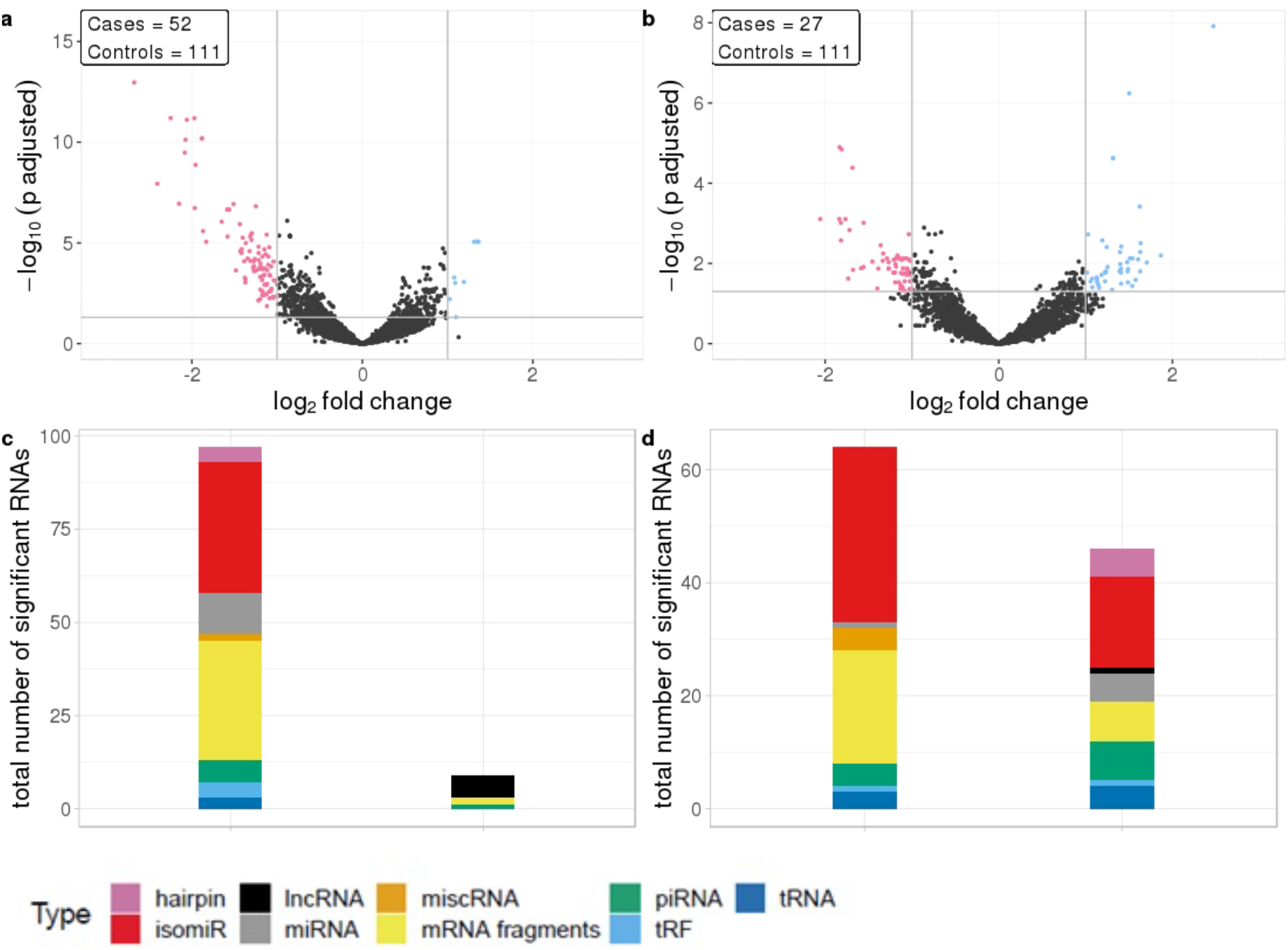
a) Differential RNA expression between seminomas and controls; b) Differential RNA expression between nonseminomas and controls; Pink represent RNAs with >1 log2 fold change reduced levels in cases and blue represent RNAs with >1 log2 fold change increased levels in cases, compared to controls. c & d) RNA class composition of elevated and reduced RNA levels with associated histology.

In contrast to TGCT and seminomas, where the differences compared to controls were generally large, nonseminomas had 63 RNAs with reduced levels and 46 RNAs with elevated levels (figure 2b), with mean log2fc values of -0.131 and 1.34 respectively. The majority of the RNAs with reduced levels consisted of mRNA fragments and isomiRs, as was also observed with all TGCT, a result driven by the seminomas. However, in nonseminomas the elevated RNAs showed higher proportions of isomiRs, and mRNA fragments, piRNAs and miRNAs as the second most abundant types (figure 2d).

### RNA signals are not associated with time to diagnosis

To investigate if there is any association between RNA signals and time to diagnosis, four different time frames during the prediagnostic period were examined. The RNA signals in all time frames exhibited similar patterns to the overall prediagnostic TGCT patterns, with the majority of significantly differentially expressed RNAs having reduced levels. This was strongest in the 2-5 years time frame with 282 RNA levels being reduced and 64 RNAs with elevated levels. The 0-2 years time frame showed the least amount of differentially expressed RNAs, with 93 RNAs showing reduced levels and 18 with elevated levels (figure 3a). Fragments containing mRNAs constitute the major proportion of the significantly changed RNAs in all time frames. However, in the 2-5 years time frame, a higher number of piRNAs (76 out of 244) had different levels, and furthermore, in the 8-10 years, the higher proportion was isomiRs (64 out of 155) (figure 3b).

**Figure 3.**
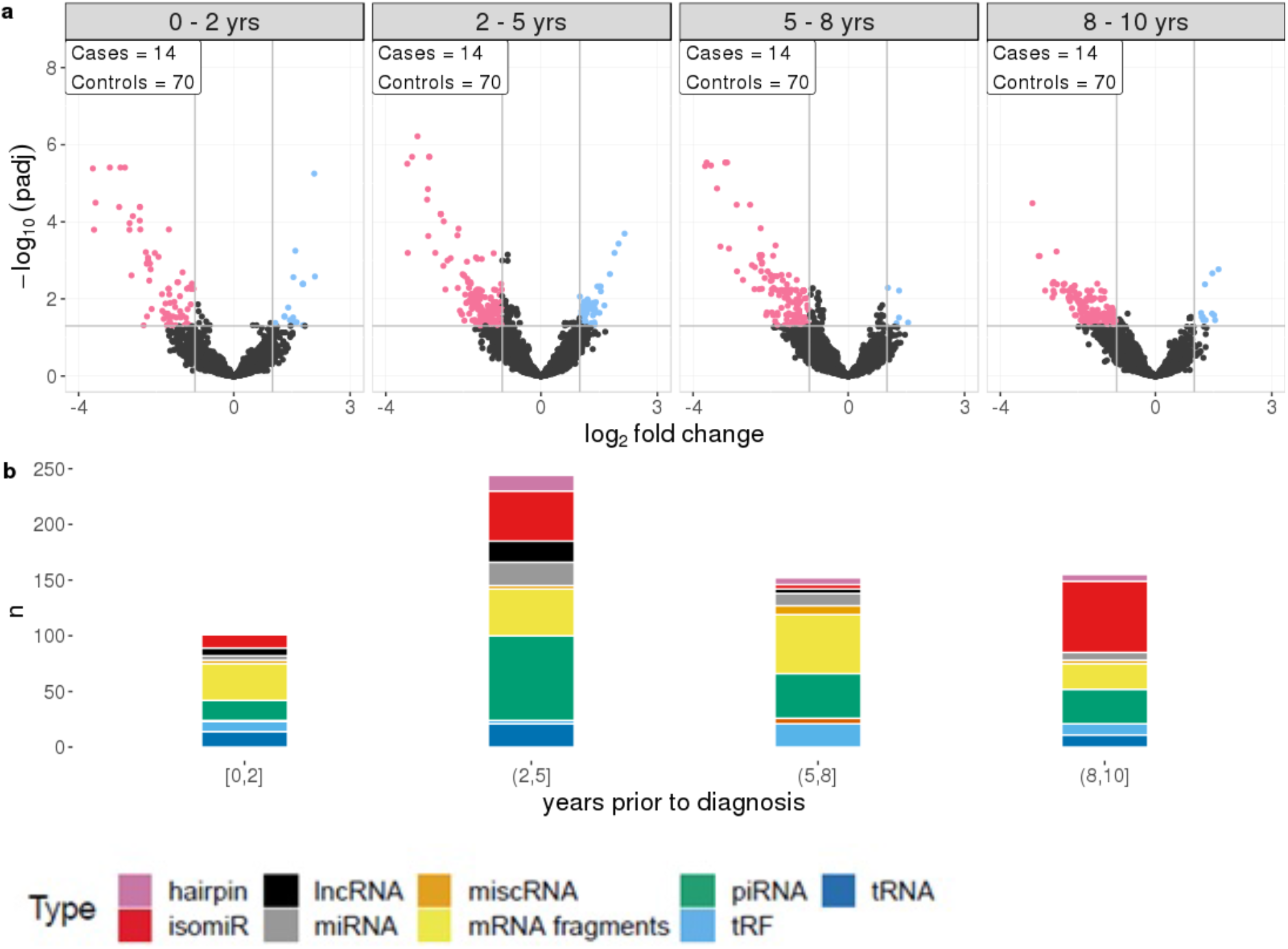
a) Differential RNA expression between cases and matched controls, across four time frames prior to diagnosis with equal sample sizes. Pink represent RNAs with >1 log2 fold change reduced levels in cases and blue represent RNAs with >1 log2 fold change increased levels in cases, compared to controls. b) RNA class composition of elevated and reduced significant RNA levels for the associated time frames.

### Specific RNA signals are independent of histology and time to diagnosis

A number of RNAs are specific or common to histologies and time frames as illustrated by the venn diagrams in figure 4. A total of 83 out of 88 of the RNAs identified were observed in both seminoma separately and all TGCT profiles. Furthermore, seminomas and nonseminomas shared 35 RNAs, and the majority of these were either mRNA fragments or isomiRs (figure 4a & c).

**Figure 4.**
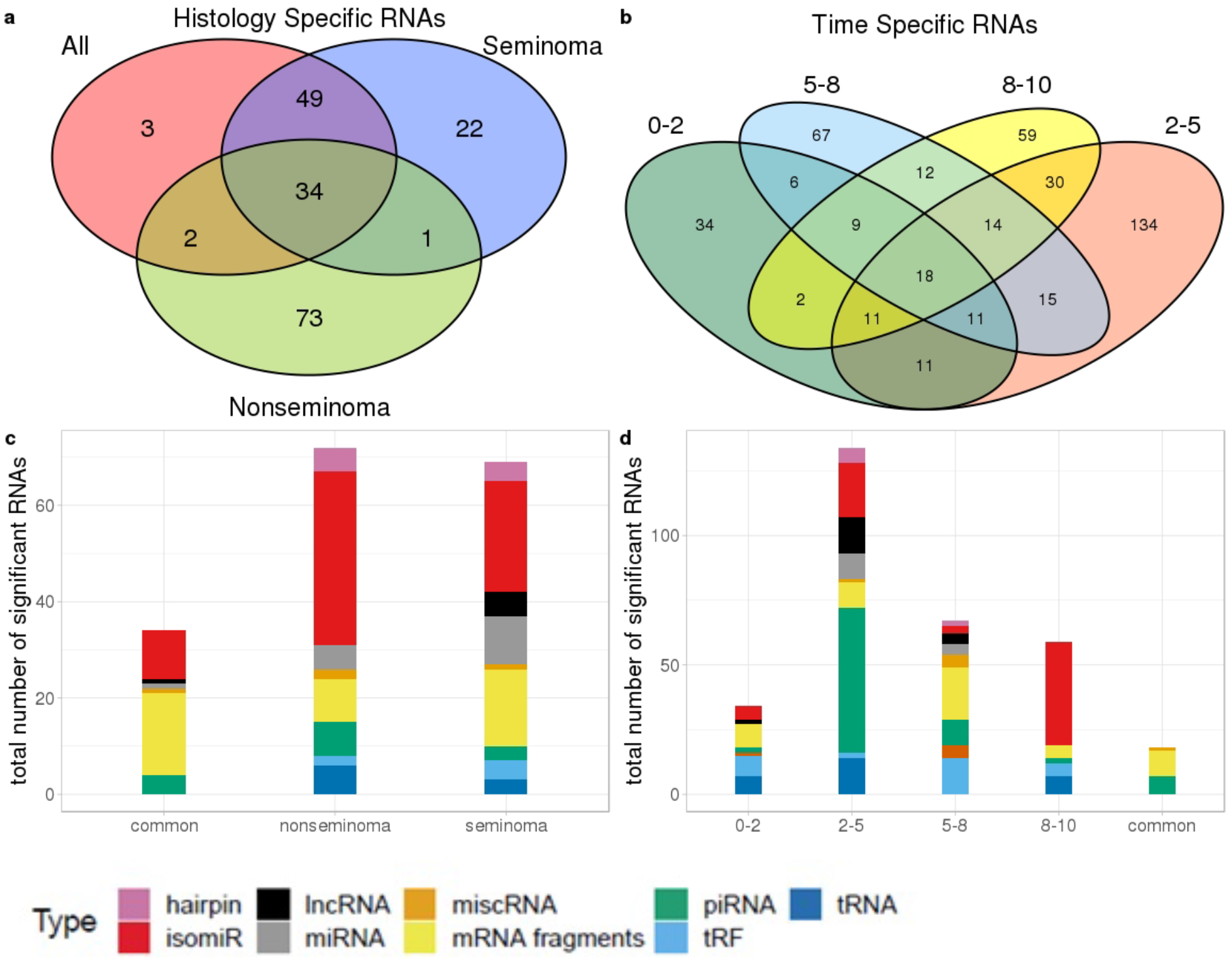
Venn diagram showing the number of a.) RNAs which are dependent and independent on histological subtype; b.) RNAs which are dependent and independent on single time periods or are observed in multiple time periods. c.) Types of RNAs differentially present in only seminomas and nonseminomas, and the 35 common types shared between both; d.) Types of differentially present RNAs unique to each time frame as well as those 18 shared between all four time frames.

An investigation into the common RNAs between time frames showed that 18 RNAs are consistently differentially expressed at all time frames, and that these RNAs were piRNAs and mRNA fragments. RNAs from the 2-5 years time frame, which has the highest number of RNAs above our cut-off, consisted of primarily piRNAs (figure 4b & d).

### mRNA fragments have consistently reduced levels in TGCT cases compared to controls

To investigate the stability of TGCT RNAs over time, we looked at nine RNAs with the lowest adjusted p values which appeared in all four time frames and their associated log2fc. Primarily, the significantly reduced RNAs were mRNA fragments which showed a relatively stable log2fc over time. BHLHE41 showed some dynamic changes across the time frames with a log2fc range between - 1.91 to -3.45, however, this could still be considered stable due to the overall log2fc remaining negative. Of all the mRNAs, a slight overall increase in log2fc was observed over time, with a mean log2fc of -2.91 at 0-2 years and -2.52 at 8-10 years. However, the lowest average log2fc appears at 2-5 years with a mean of -3.08, followed by a mean of -3.06 at 5-8 years. This showed that during the extended time frame of 0-8 years, there was a stability in the RNA levels, with a slight increase at 8-10 years (figure 5).

**Figure 5.**
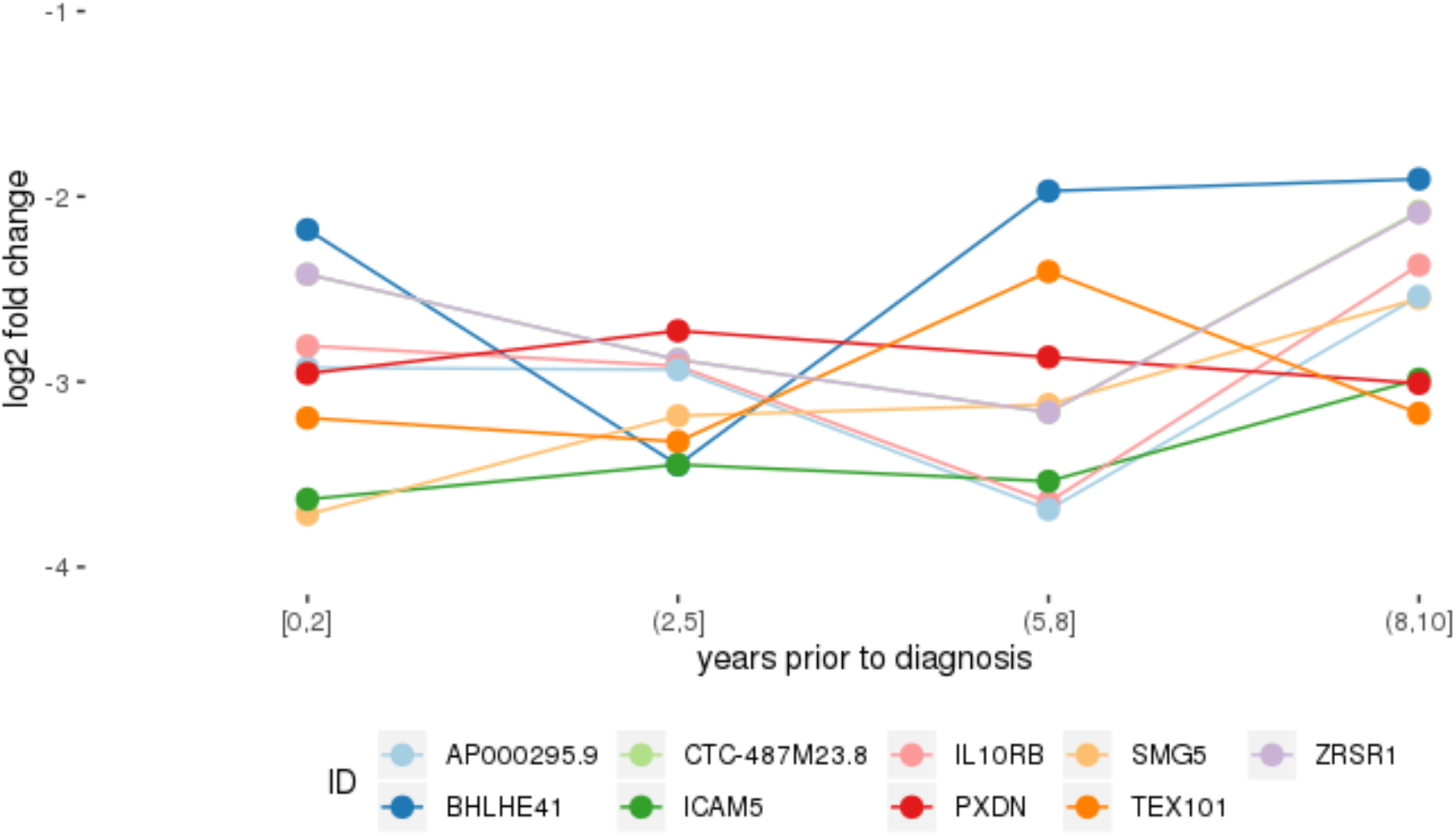
Log2 fold change of RNAs that had significantly stably changed levels between cases and controls in all four time periods.

### Common cancer related pathways were enriched in specific time periods and histologies

Enrichment analysis showed multiple pathways enriched for significant miRNA targets and mRNA fragments. Of the significant pathways, only one (Axon guidance [mean p adjusted = 0.0094]) appeared in all seven subgroups, and two appeared in six of the seven sub-analyses (Ras signalling pathway [mean padj =0,10], MAPK signalling pathway [mean padj =0,043]). In total there are 19 pathways that span at least 3 of the time frames, with the remaining 6 only appearing in two of the four. Nonseminoma and 0-2 year time frame had the lowest number of significant enriched pathways with only 3 pathways in each.

Looking at each sub analysis, enrichment of miRNAs and mRNA fragments among the 88 significant RNAs in all TGCTs revealed several cancer related pathways, including mTOR, MAPK and ErbB2 (figure 6). In seminomas the expressed miRNAs and mRNA fragments also showed the cancer related pathways mTOR, MAPK and AMPK as well as the proteoglycans in cancer pathway. Similar pathways were present in nonseminomas as well, with miRNAs and mRNA fragments showing cancer related pathways, with mTOR, MAPK, ERBb2 and PI3K-Akt showing highest significance (figure 6). Finally, all four time frames showed significant pathways, including the mTOR signalling pathway which was one of the top 10 more significant pathways throughout, with the MAPK and Erb2 pathways also showing up once more within the time frames (figure 6).

**Figure 6.**
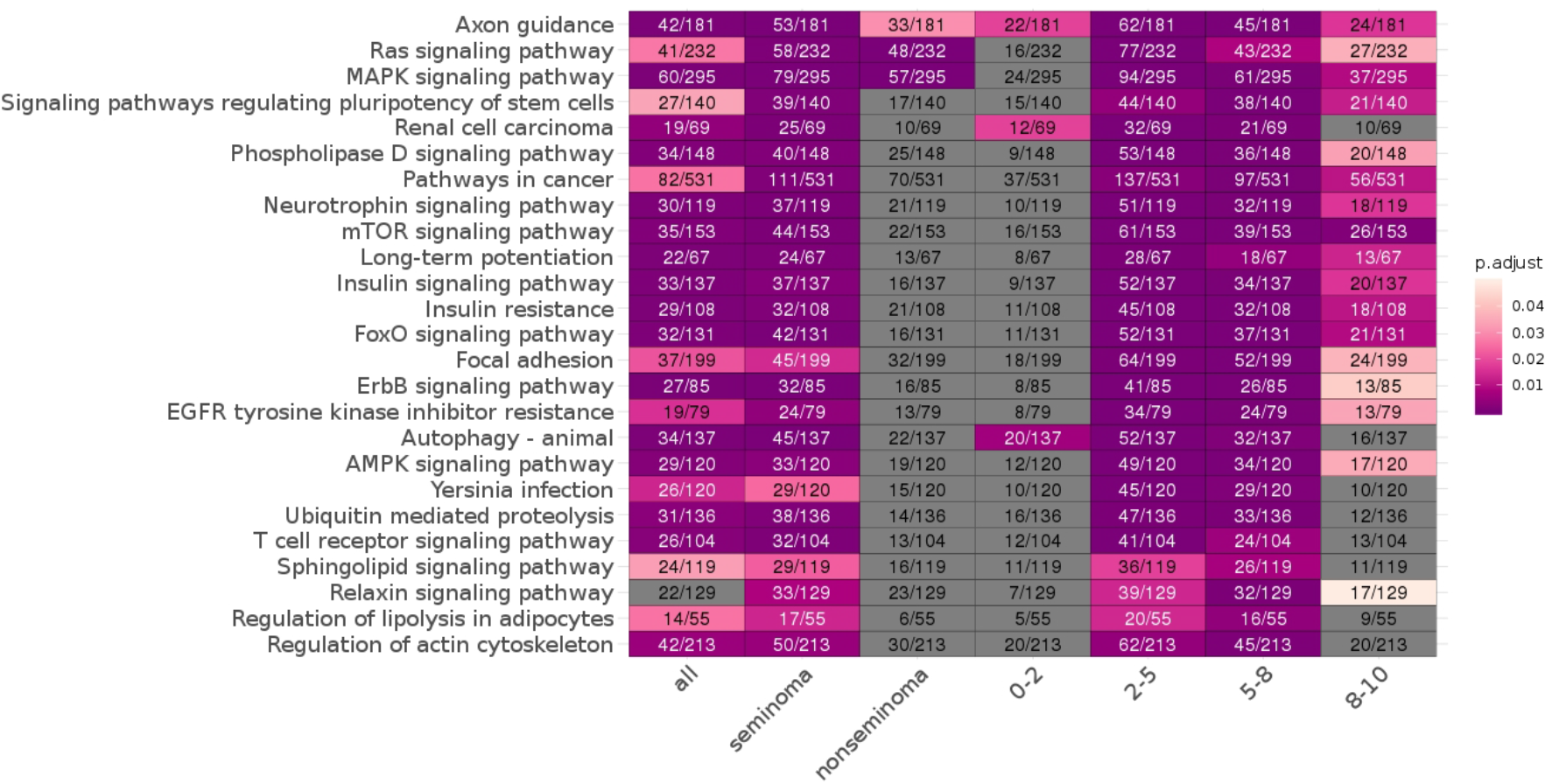
KEGG pathways enriched for differentially expressed mRNAs and miRNA targets for all TGCT vs controls, seminoma vs controls, nonseminomas vs controls, 0-2, 2-5, 5-8 and 8-10 years prior to diagnosis vs controls are shown. The pathways are colour coded according to p values and the number of enriched genes and total number of genes in the pathways are also shown. The 25 most commonly enriched pathways for these analyses were selected for presentational purposes

## Discussion

We found the RNA profiles in pre-diagnostic serum of TGCT patients to be different from that of controls, and observed that the difference was relatively stable across the pre-diagnostic time period of 10 years. Although previous studies have identified TGCT specific RNA profiles in patients [Palmer et al., 2010; Murray et al., 2011; Gillis et al., 2007], this has not been described for the pre-diagnostic period, as seen in our study. In contrast to our findings, highly dynamic pre-diagnostic RNA signals have been observed in lung cancer and breast cancer samples [Umu et al., 2020; Lund et al., 2016; Holden et al., 2017]. The reason for this contrast may partly be due to the localization of the tumour within the testis. Typically, the developed GCNIS is found above the basement membrane in seminiferous tubules, which are located in the intratesticular environment and therefore separate from the bloodstream via the blood testis barrier (BTB) [Spiller and Bowles, 2017; Reuter, 2005]. The BTB’s primary function is to prevent both antibodies and T lymphocytes from affecting the testis [Bart et al., 2002]. Furthermore, angiogenesis in breast and lung cancer is well documented, showing a closer physical interaction between these tumours and circulatory system than what applies to TGCT [Herbst et al., 2005; Longatto et al., 2010]. GCNIS may also be less affected by the immune system [Endo and Inoue, 2019; Weis and Cheresh, 2013] as the BTB provides adequate protection against immune response during the early stages of the malignant growth. Another potential implication of the presence of the BTB function is a delay in serum signals. Changes in the GCNIS embedded in the seminiferous tubules do not necessarily appear in blood unless the tumour causes disruption of the BTB. [Cheng and Mruk, 2012]. Further studies are needed to show how the interference of TGCT with the BTB affects the levels of circulating RNAs.

In this study we found that differentially expressed levels of mRNAs were related to both testis development and known cancer pathways, as well as the lncRNA, TSIX which exhibited increased levels in TGCT patients. The role of TSIX in X-chromosome inactivation (XCI) has been widely reported [Del Rosario et al., 2017; Gayen et al., 2015; Turner et al., 2002], acting as the antisense repressor of XCI. The expression of TSIX is limited to pluripotent cells and the testis. A zinc finger protein, GLI, binds to the 5’ ends of TSIX to reduce TSIX expression and block the initiation of XCI. Physiologically, XCI is observed during spermatogenesis, but can also be expressed in TGCTs through supernumerary X chromosome constitution [Lobo et al., 2019]. XCI has been observed in TGCT of different histogenesis and may be a potential biomarker for some types of TGCT [Looijenga et al., 1997].

Alongside increased levels of TSIX, we also observed a positive log2fc with RAB21, an oncogene involved in mediating endocytosis and connected to the tumorigenesis and vesicle transport mechanisms of the other Rab proteins [Tzeng and Wang, 2016]. The silencing of RAB21 has also been utilised to induce apoptosis in glioma cells and can also be used to significantly inhibit cell growth [Ge et al. 2017]. The increased presence of RAB21 in the serum of patients after diagnosis of TGCT has not been observed before, however, previous studies have elucidated the function of a similar RAS family associated protein, RAB12, in rat testis development, especially noting the high expression in Sertoli cells (SC) [Iida et al., 2005]. RAB GTPases have been identified as mediators for vesicle trafficking in cancer, including both RAB21 and RAB12 [Tzeng et al., 2016].

Investigating of the different histology patterns showed that despite the increased positive log2fc in nonseminomas, there are still RNA signals, such as mRNA TEX101, that are independent of the histology. TEX101 is involved in the production of spermatozoa, and studies using animal models have shown that males with a disrupted TEX101 gene often produce spermatozoa that are unable to fertilise, despite looking normal [Fujihara et al., 2013]. Previous studies have found links between TGCT and changes in fertility [Skakkebæk et al., 2001]. These findings include men with TGCT having fewer children than average, as well as a lower proportion of those children being male. Other studies have found that abnormal semen characteristics can be observed in men who later develop TGCT, indicating that these two aspects are aetiologically linked [Jacobsen et al., 2000]

As hypothesised previously, the functions of the mRNAs identified as stable across all time frames could give some indication as to how GCNIS develops into tumours. Fragments of the mRNA for AP000295.1 had lower levels in cases compared to controls. AP000295.1 mRNA expression has been previously noted in the Human Protein Atlas (HPA) project, and HPA shows RNA-seq tissue data with high protein-coding transcripts per million (pTPM) counts of AP000295.1 in the blood constituents, granulocytes and monocytes [Uhlen et al., 2015]. AP000295.1 belongs to a network of genes associated with kinase binding and type I interferon binding, AP000295.1 is also a paralog of the gene IFNAR2 with 100% match between target and query genes as seen using Ensembl (release 99) [Cunningham et al., 2019]. Of note here is that IFNAR2 has been detected on the surface of pre-Sertoli cells (pSC) [Edgar et al., 2013]. Previous studies have demonstrated the importance of Sertoli cells in the BTB formation and homeostasis [Gerber et al., 2016], thus potentially explaining the presence of an RNA signal in serum despite the impermeable BTB. Through the reduction in expression of IFNAR2 in pSC’s surface, inadequate formation and maintenance of the BTB could occur, causing a breakdown of this filter between the testis and the blood, thus explaining higher presence of signals of tumour development in serum.

Circulating miRNAs have been identified as both prognostic and diagnostic markers in biliary tract cancers (BTC), and this has led to earlier diagnosis and less invasive procedure of BTC than previously used techniques [Letelier, 2016]. The circulating miRNAs used for BTC diagnosis were reported to be stable in serum and showed significantly different miRNA expression in patients with BTC and controls, and there also may be ethnic differences. Circulating biomarkers has also been identified in colorectal cancer (CRC), specifically the exosomal miRNAs, miR-150-5p and miR-99b-5p were found to be downregulated in CRC patients compared to healthy individuals [Zhao et al., 2019]. This demonstrates the potential for their later use in clinical settings as a non-invasive diagnostic technique.

Pathways enrichment analysis showed overall many associated cancer related pathways across the 10-year period prior to TGCT diagnosis. The time frames 2-5 and 5-8 years prior to diagnosis showed the highest numbers of enriched pathways, many of which were cancer related. A possible explanation for the absence of enriched pathways in the 0-2 year time interval, could be an overall increase of all RNA types in serum as the tumour develops, this would effectively mask cancer pathways from being detected due to the large amounts of noise from aberrant RNA expression.

A strength of this study is the large and robust RNA dataset. To our knowledge this is the largest study of the RNA profiles from TGCT patients prior to diagnosis also including harmonized confounders collected from health surveys [Hjerkind, 2017]. The time interval throughout which the samples were collected is also a strength of this study. With up to 10 years time before diagnosis we were able to observe TGCT development and elucidate the body’s response.

The sample size is still a limitation, and stratified analyses on histology and time to diagnosis could benefit from an increase in statistical power. Serum samples and survey data were collected over a period of more than 30 years, and lifestyle factors such as smoking and BMI have changed within this time period as well as carcinogens in the environment, such as organochlorine pesticides [Irigaray et al., 2007; Belpomme et al., 2007; McGlynn and Trabet, 2012]. This was partially controlled for by matching.

Primarily, the new insights into TGCT carcinogenesis helps us better understand the development of the disease, which is thought to be initiated *in utero*. Stable serum RNAs in the pre-diagnostic period have the potential to be biomarkers for earlier detection. Early diagnosis of TGCT would lead to less use of cisplatin treatment, and thereby reduce long-term adverse effects, such as risk of second primary cancer and cardiovascular disease. Hazard ratios for all secondary cancers after a single cisplatin-based chemotherapy cycle were significantly lower than hazard ratios after two or more cycles [Hellesnes et al., 2019].

In conclusion, RNA levels associated with cancer related pathways were different between individuals who developed TGCT, compared to matched controls. There is some loss of pathway signals closer to diagnosis, however, there is inherent reduced levels of serum RNAs in cases compared to controls for most of the 10 year pre-diagnostic follow-up time. The presence of these stable RNA signals may help in the identification of biomarkers give insights to molecular mechanisms driving TGCT development.

## Data Availability

The datasets generated and analysed during the current study are not publicly available since individual privacy could be compromised but are available from the corresponding author on reasonable request and with appropriate approvals.

## Acknowledgements

We would like to acknowledge Cecilie Bucher-Johannessen, Marianne Lauritzen, Kari Furu and Magnus Leithaug for performing laboratory and coordination tasks. We acknowledge the Norwegian Institute of Public Health for access to survey data in this study. The sequencing service was provided by the Norwegian Sequencing Centre (http://www.sequencing.uio.no), a national technology platform hosted by Oslo University Hospital and the University of Oslo supported by the Research Council of Norway and the Southeastern Regional Health Authority.

